# Whole-genome sequencing analysis of clozapine-induced myocarditis

**DOI:** 10.1101/2021.07.26.21261157

**Authors:** Ankita Narang, Paul Lacaze, Kathlyn J Ronaldson, John J McNeil, Mahesh Jayaram, Naveen Thomas, Rory Sellmer, David Crockford, Robert Stowe, Steven C. Greenway, Christos Pantelis, Chad A Bousman

**Affiliations:** Department of Medical Genetics, University of Calgary, Calgary, AB, Canada; Department of Epidemiology and Preventive Medicine, Monash University, Melbourne, VIC, Australia; Adult Mental Health Rehabilitation Unit, North Western Mental Health, Melbourne Health, Western Health, Sunshine Hospital, St Albans, VIC, Australia; Melbourne Neuropsychiatry Centre, Department of Psychiatry, The University of Melbourne and Melbourne Health, Western Centre for Health & Education, St Albans, VIC, Australia; Department of Psychiatry, University of Calgary, Calgary, AB, Canada; Departments of Psychiatry and Neurology (Medicine), Neuropsychiatry Program, and Djavad Mowafaghian Centre for Brain Health, University of British Columbia, Vancouver, BC, Canada; Department of Pediatrics, Cumming School of Medicine, University of Calgary, Calgary, AB, Canada; Department of Biochemistry & Molecular Biology, Cumming School of Medicine, University of Calgary, Calgary, AB, Canada; Alberta Children’s Hospital Research Institute, Cumming School of Medicine, University of Calgary, Calgary, AB, Canada; Department of Cardiac Sciences, Cumming School of Medicine, University of Calgary, Calgary, AB, Canada; Libin Cardiovascular Institute, Cumming School of Medicine, University of Calgary, Calgary, AB, Canada; Department of Physiology & Pharmacology, University of Calgary, Calgary, AB, Canada; Hotchkiss Brain Institute, Cumming School of Medicine, University of Calgary, Calgary, AB, Canada

## Abstract

One of the concerns limiting the use of clozapine in schizophrenia treatment is the risk of rare but potentially fatal myocarditis. Our previous genome-wide association study and human leucocyte antigen analyses identified putative loci associated with clozapine-induced myocarditis. However, the contribution of DNA variation in cytochrome P450 genes, copy number variants and rare deleterious variants have not been investigated. We explored these unexplored classes of DNA variation using whole-genome sequencing data from 25 cases with clozapine-induced myocarditis and 25 demographically-matched clozapine-tolerant control subjects. We identified 15 genes based on rare variant gene-burden analysis (*MLLT6, CADPS, TACC2, L3MBTL4, NPY, SLC25A21, PARVB, GPR179, ACAD9, NOL8, C5orf33, FAM127A, AFDN, SLC6A11, PXDN)* nominally associated (p<0.05) with clozapine-induced myocarditis. Of these genes, 13 were expressed in human myocardial tissue. Although independent replication of these findings is required, our study provides preliminary insights into the potential role of rare genetic variants in susceptibility to clozapine-induced myocarditis.

## INTRODUCTON

Clozapine initiation for patients suffering from treatment-resistant schizophrenia is recommended at the earliest possible opportunity by clinical guidelines,^1, 2^ and there is evidence for its superior efficacy in this difficult-to-treat population compared with other pharmacological treatments^3^. Regulatory requirements around serious adverse event monitoring have largely addressed the risk of undiagnosed agranulocytosis. However, there has been ongoing concern about a serious idiosyncratic adverse drug reaction (ADR) involving inflammation of the myocardium.

In 1999, Kilian and colleagues^4^ reported the rate of myocarditis (both fatal and non-fatal) as 1.9 per 1000 patients starting clozapine. However, more recent projections suggest rates as high as 3% in the context of systematic monitoring, with fatality rates ranging from 7% to 75%^5^. Despite these alarming rates, our current ability to identify those at greatest risk for clozapine-induced myocarditis is inadequate. This uncertainty has led to reduced clozapine prescription and arguably poorer patient outcomes. Although research to uncover risk factors has identified a few clinical markers^6,7^, these factors do not account for all the risk. As such, identification of additional markers is needed to reduce or prevent fatalities and morbidity resulting from clozapine-induced myocarditis, potentially facilitating expanded and safer use of this highly effective medication.

Genomic analysis is a promising and largely unexplored approach for identifying those at risk of idiosyncratic ADRs. In fact, our recent genome-wide association study (GWAS) and human leucocyte antigen (HLA) analyses of clozapine-induced myocarditis identified four independent loci (*SIX3, EFHC1, ADAM7*, and *GNA15*) suggestive of increased myocarditis risk (P < 1 × 10^−6^), with odds ratios ranging from 5.5 to 13.7. We also found that the HLA-C*07:01 allele was associated with 2.89-fold (95% CI: 1.11–7.53) greater odds of clozapine-induced myocarditis^8^. To build on these promising findings, we undertook the first whole-genome sequencing study to examine different classes of DNA variation not measured in our previous genetic study, including rare deleterious coding variants, copy number variants, and candidate variation in pharmacogenes associated with clozapine metabolism. Herein, we report our results among a cohort of individuals with and without a history of clozapine-induced myocarditis, in collaboration with members of the Pharmacogenomics of Clozapine-Induced Myocarditis (PROCLAIM) Consortium^9^.

## METHODS

### Study Participants

Participants with (n = 25) and without (n = 25) a history of clozapine-induced myocarditis were recruited by investigators affiliated with the PROCLAIM Consortium at the University of Calgary, University of Melbourne, and Monash University. For this study, we included 34 participants (19 cases and 19 controls) included in our previous GWAS study^8^. For each recruited case of clozapine-induced myocarditis, an age (± 5 years), sex, and ancestry-matched clozapine-tolerant control was recruited from the same site. Genetic ancestry of controls and cases was further assessed by principal component analysis using the smartpca module of the EIGENSOFT package (Supplementary Figure S1). Individuals were eligible for inclusion into the study if they: (a) were aged 18 to 65 years, (b) had a clinical diagnosis of a schizophrenia-spectrum disorder, (c) had accessible medical records, and (d) had a history of clozapine therapy. For cases, an additional inclusion criterion was a history of myocarditis evidenced by abnormal troponin I/T levels (>2 times upper limit of normal) or signs of left ventricular dysfunction with or without elevated C-reactive protein (CRP) levels (>100mg/L) during the first 30 days of clozapine therapy, which aligns with current monitoring guidelines^10^. An additional inclusion criterion for clozapine-tolerant controls was evidence of clozapine therapy for a minimum of 45 days with no documented history or clinical suspicion of myocarditis. The present study was approved by the Melbourne Health Human Research Ethics Committee and University of Calgary Conjoint Health and Research Ethics Board. The study also complied with the Declaration of Helsinki and its subsequent revisions. All participants or next of kin provided written informed consent prior to participation.

### Whole genome sequencing

Genomic DNA (750 – 1000 ng, A260/A280 ratio > 1.8) was isolated from whole blood samples using standard procedures and shipped to the Centre for Applied Genomics at the Hospital for Sick Children (Toronto, Canada) for library preparation and sequencing. Libraries were prepared for all samples using the Illumina TruSeq PCR-free DNA library preparation kit. Each library was subjected to 150 base, paired end sequencing on three lanes of the Illumina HiSeqX.

The Illumina DRAGEN (version 07.021.382.3.4.9, Dynamic Read Analysis for GENomics) Bio-IT platform was used for aligning reads to the human genome (reference build hg19). After alignment, sorting and duplicate marking was performed to generate aligned BAM files. Next, sample-wise gVCF files were generated using DRAGEN’s haplotype calling algorithm and then these files were run through the joint calling algorithm to generate a joint VCF file. dbSNP (v150) was used to annotate variants. The jointly called VCF file was further recalibrated using the Variant Quality Score Recalibration (VQSR) algorithm to produce a high-quality variant dataset. The VQSR model was trained for single nucleotide polymorphisms (SNPs) and insertion/deletions (indels) using high quality variant data from different resources (see Supplementary Methods for more details). For SNPs, the VQSR model was trained using data from HapMap3.3, 1000 Genomes Project (Phase 1 SNP sites and Omni 2.5 M SNP array) and dbSNP (v150), with a sensitivity threshold of 99.5%. For indels, the VQSR was trained using data in the 1000 Genomes Project (gold standard sites) and Illumina Platinum sites, with a sensitivity threshold of 99.0%. Only “PASS” calls were annotated using ANNOVAR after VQSR implementation.

### Pharmacogenetic variant analysis

The Stargazer v1.08^11^ tool was used to call diplotypes, calculate activity scores, and assign genotype-inferred metabolizer phenotypes for seven genes (*CYP1A2, CYP2C9, CYP2C19, CYP2D6, CYP3A4, CYP3A5, CYP2E1*) relevant to clozapine metabolism^12^. Stargazer uses whole genome sequencing data to find phased haplotypes comprised of SNPs, small indels, and large structural variants, and then maps these haplotypes to star alleles based on translation tables maintained by the Pharmcogene Variation Consortium^13^. Notably, previous work has shown Stargazer recalled 100% of star alleles present in the Genetic Testing Reference Material Coordination Program’s consensus genotypes^11^. Furthermore, a comparison of Stargazer-predicted diplotypes for the seven target pharmcogenes with the diplotypes generated by the PharmacoScan Solution (Thermofisher, Carlsbad, USA) array in a subset of our sample (n = 22) tested on both platforms found high diplotype (94.8%) and phenotype (98.1%) concordance (Supplementary Table S1).

Prior to analysis, activity scores for the seven target pharmacogenes were corrected for concomitant inhibitors or inducers (Supplementary Table S2) using a method previously described^14^. In brief, activity scores for one or more of the seven genes were multiplied by zero if a strong inhibitor of the corresponding enzyme was present. For moderate inhibitors the activity score was multiplied by 0.5 and for inducers the activity score was multiplied by 1.5. In cases where both an inhibitor and inducer of the same enzyme were present, the uncorrected activity score for the corresponding gene was retained, as consensus on how to correct activity scores in this situation has not been established. The corrected activity scores were then used to assign a corrected metabolizer phenotype for each of the seven target genes. To determine differences in activity scores and metabolizer phenotypes between the cases and controls, we used the Mann-Whitney U and chi-square tests, respectively. P values were corrected for multiple testing using the Benjamini–Hochberg (B–H) step-up procedure^15^.

### Copy number variant (CNV) analysis

CNVs (variants with length ≥1kb) were called using the DRAGEN algorithm and self-normalization method. A merged CNV file was created by compiling copy number state and segmentation mean values for all samples. Segmentation mean values were log transformed to construct copy number variable regions (CNVRs). Ten regions where segmentation mean and log transformed segmentation values had conflicting CNV status were excluded (Supplementary Table S3). We used two different tools - CNVRuler^16^ and CoNVaQ^17^ to construct CNVRs and perform CNVR association analyses. Using CNVRuler, logistic regression was performed using the first two principal components as covariates, and a minor allele frequency threshold of greater than 0.1. CNVRuler performs multiple testing using the B–H step-up procedure^15^. CNVR association analysis was also performed using CoNVaQ with permutation (n=4000) testing by swapping phenotype labels. CNVR encompassing genes were retrieved from the UCSC Genome Browser.

### Gene burden analysis and functional annotation of candidate genes

We performed burden analysis using rare and deleterious variants, defined based on the minor allele frequency from global databases and combined annotation dependent depletion (CADD) annotation score using ANNOVAR software^18^. A variant was considered rare if the maximum allele frequency was less than 0.05 in the 1000 Genomes Project^19^, ESP6500^20^, ExAC^21^, CG46^22^ or gnomAD databases^23^. A deleterious variant was defined by a CADD Phred score greater than 20. We used CADD scores to include functionally important variants from protein-coding and non-coding regions. Variants that met both criteria were extracted using BCFtools (v1.9)^24^ and burden analysis was performed on these selected variants. Two different methods were used - Combined and Multivariate Collapsing (CMC) and the sequencing kernel association test (SKAT-O), both implemented in RVTESTS^25^. Permutation analysis (10,000 iterations) was carried out in R by permuting phenotype labels (case and control) of observations (i.e. total allele counts) to derive empirical p-values. We also used TRAPD (Testing Rare vAriants using Public Data)^26^ software to compute the frequency of rare and deleterious alleles of prioritised genes using genome data from 15,708 individuals in the gnomAD v2 database (build hg19)^23^ and then compared these frequencies with those observed in our cases and controls using a two proportion Z-test. P-values were corrected using the Bonferroni method.

To evaluate the tolerance of genes for functional genetic variation, Residual Variation Intolerance Score (RVIS) scores were fetched using the web-based tool Gene Intolerance^27^. RVIS scores are used to rank genes in terms of their functional tolerance^28^. Genes with negative scores represent less tolerance to functional variations. Gene enrichment analysis was performed using ToppFun^29^. Finally, we assessed the mRNA expression of candidate genes in left ventricular-septal tissue from 21 healthy adult donors using published RNASeq data^30^. Raw RNASeq data was retrieved from the Zenodo Data repository^31^. Raw read counts were normalized with DESeq2, using the median of ratios method^32^.

## RESULTS

A summary of the study design and key results are shown in Figure 1.

**Figure 1.**
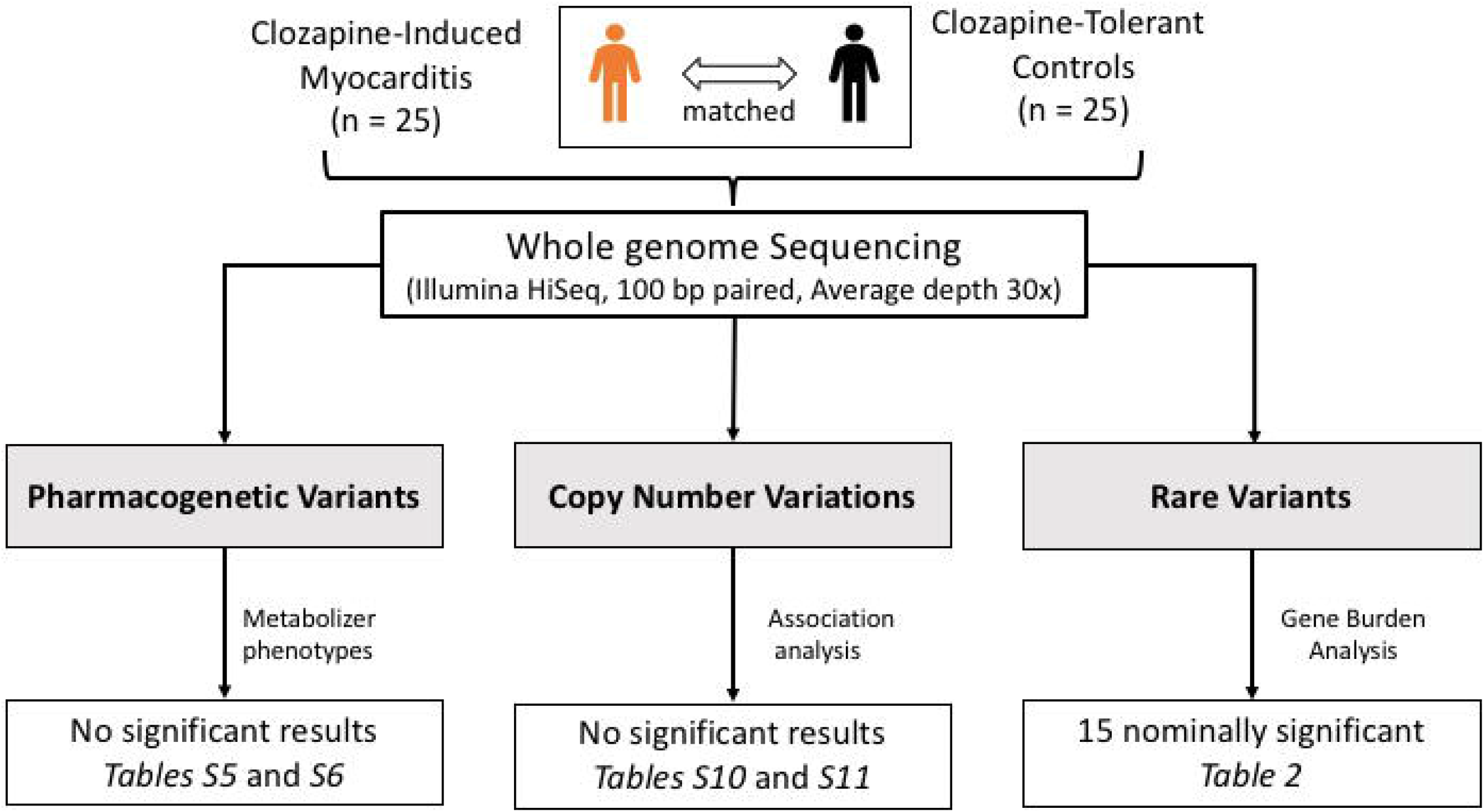
PROCLAIM study design, analysis and key results. Age, sex and ethnicity matched treatment resistant schizophrenia patients who did (cases) or did not (controls) develop myocarditis following clozapine exposure were recruited. Using whole genome sequencing data, candidate pharmacogenetic variants, copy number variable regions and rare variants were examined for their association with clozapine-induced myocarditis.

### Study participants and sequencing

Participant characteristics and relevant clinical information for the two patient groups are shown in Table 1. Cases and controls did not differ significantly in their demographic or clinical characteristics, with the exception of titration slope. On average, cases received a faster titration than controls (p = 0.006). However, *post hoc* comparisons of case and control titration slopes with those recommended in titration protocols used in Canada^33^, Australia^34^, and United Kingdom^35^ showed that neither case nor control slopes differed from these national recommendations (Figure 2). In addition, *post hoc* examination showed no difference in titration slopes by ancestry (Supplementary Figure S2).

**Table 1.**
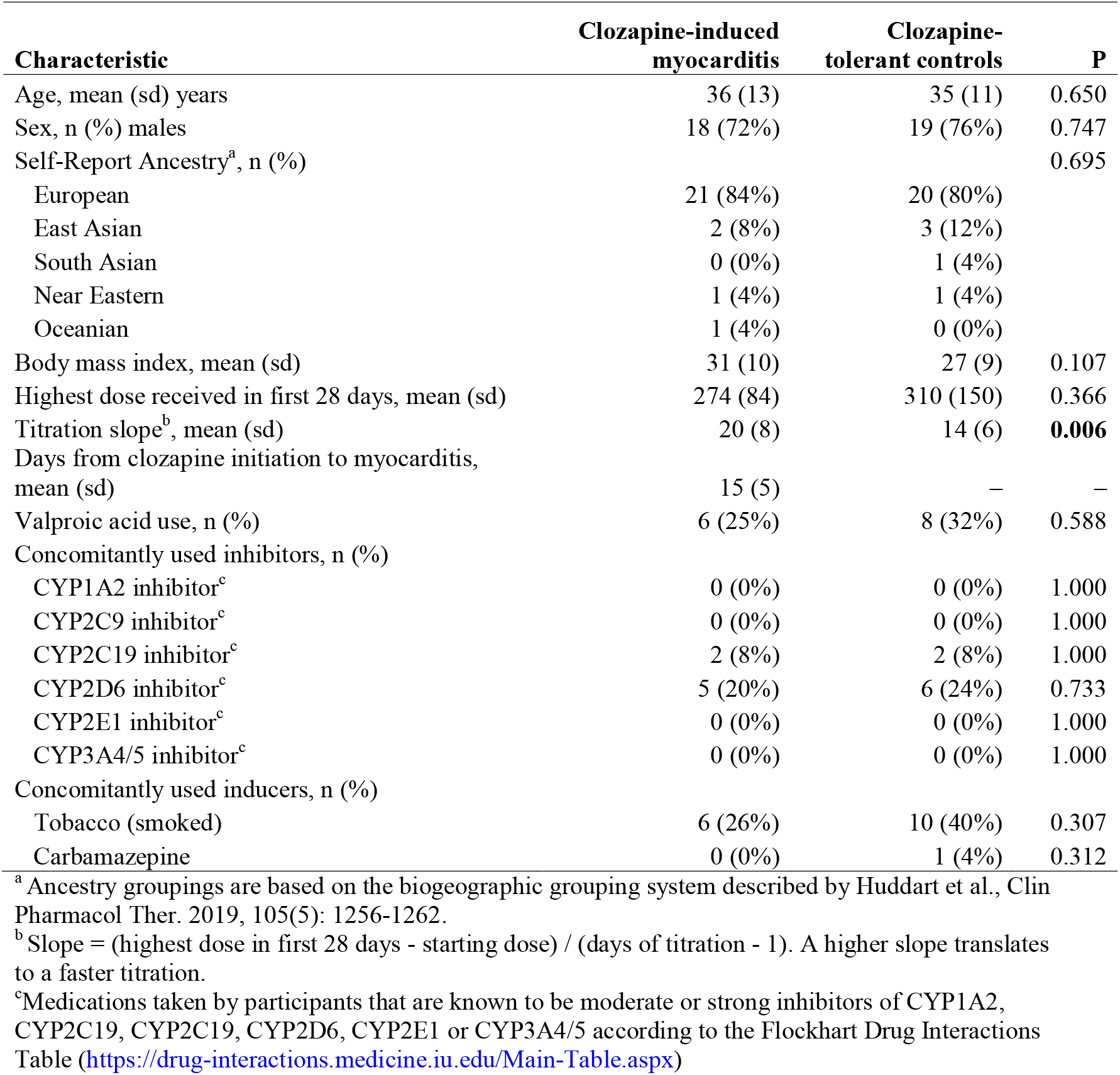
Demographic and clinical characteristics of participants with (N = 25) and without (N = 25) a history of clozapine-induced myocarditis

**Figure 2.**
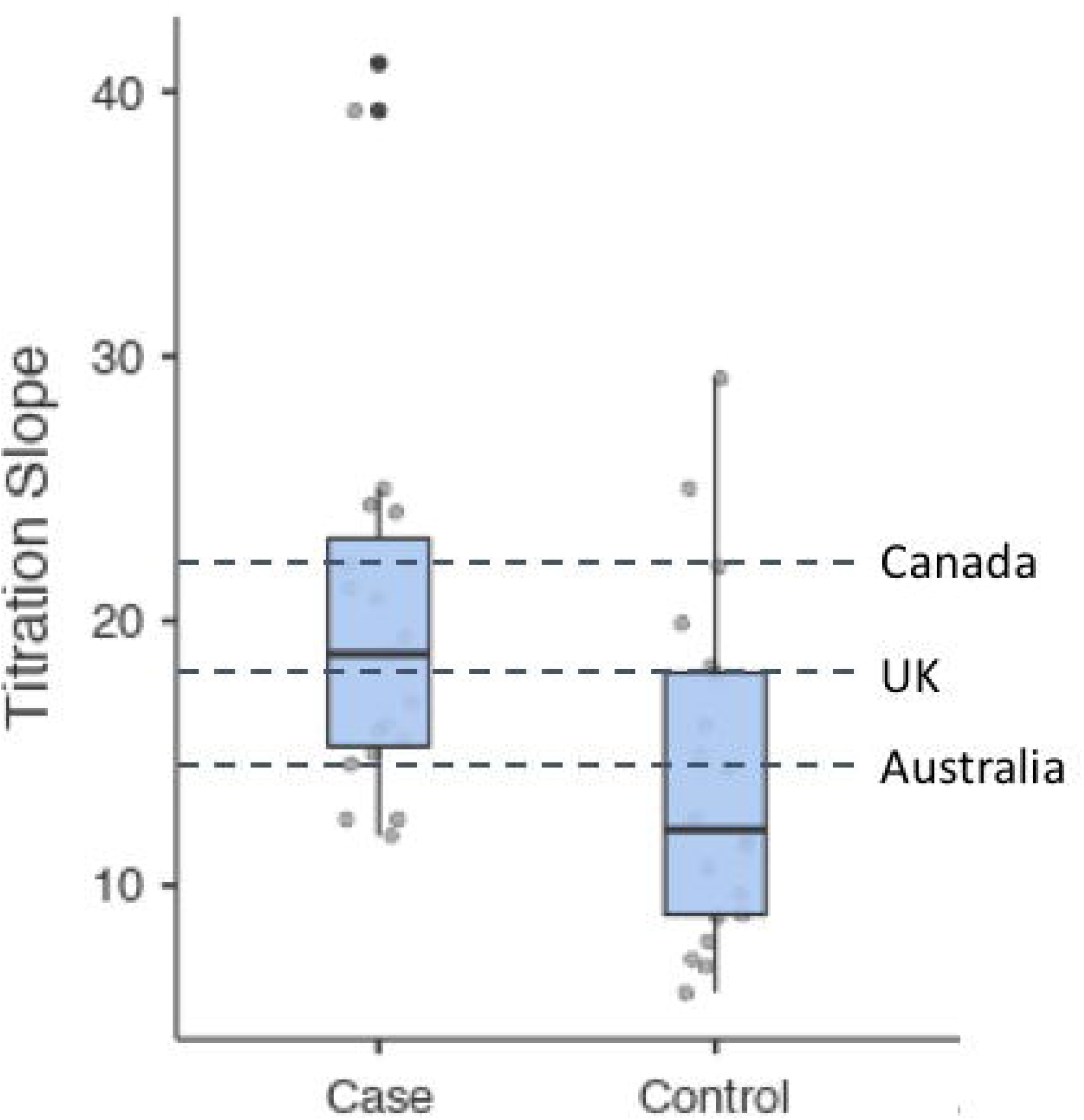
Distribution of titration slopes by history of clozapine-induced myocarditis. Cases differed significantly from controls (p = 0.006). Dashed lines represent titration slopes recommended by clozapine dosing protocols in Canada, United Kingdom, and Australia. Case titration slopes did not differ from slopes recommended by protocols used in Canada (p = 0.544, United Kingdom (p = 0.795), or Australia (p = 0.259). Likewise, control titration slopes did not differ from slopes recommended by protocols used in Canada (p = 0.187, United Kingdom (p = 0.248), or Australia (p = 0.869).

One of the control samples failed initial quality assessment post-sequencing, resulting in 25 cases and 24 controls available for further analysis with an average depth of reads and average read mapping across the samples of 33.65x and 92.9%, respectively (Supplementary Figure S3). Ti/Tv (transition/transversion) ratio for both raw (2.00) and filtered (2.02) calls were in the acceptable whole genome range. On average, there were 3,833,700 SNPs, 953,459 indels and 619 CNVs per individual (Supplementary Table S4). The final jointly called dataset contained 16,014,407 variants (both SNPs and indels).

### Pharmacogenetic variants

Observed diplotype frequencies for the seven targeted pharmacogenes, along with the corresponding uncorrected activity scores and genotype-inferred metabolizer phenotypes among cases and controls, are shown in Supplementary Table S5. No significant differences in uncorrected or corrected activity scores were found between cases and controls (Supplementary Table S6) nor were differences detected in the distribution of genotype-inferred metabolizer phenotypes (Supplementary Table S7). Furthermore, titration slopes were not correlated with activity scores for the seven targeted pharmacogenes in cases or controls (Supplementary Table S8). Importantly, *post hoc* gene burden analysis did not reveal any differences in enrichment of rare and deleterious variants in the seven genes (*CYP1A2, CYP2C9, CYP2C19, CYP2D6, CYP3A4, CYP3A5, CYP2E1*) known to be involved in clozapine metabolism.

### Copy number variants

We identified 30,340 CNVs across the 49 samples. Comparing the average number of CNVs between cases and controls revealed no statistically significant differences in losses (cases: mean = 498, ± standard deviation = 28; controls: 503 ± 19, p = 0.42) or gains (cases: 102 ± 10 vs. controls: 117 ± 8, p = 0.26) (Supplementary Table S9). We also did not detect significant differences with respect to the average size of the losses (cases: 10.54 ± 61.6 kb vs. controls: 10.57 ± 59.5 kb, p = 0.97) or gains (cases: mean 141.37 kb, sd = 721.9 kb; controls: mean = 134.01 kb, sd = 533.5 kb, p = 0.66) (Supplementary Table S9). The CNVRuler constructed 3,178 copy number variable regions (CNVRs) among which five were found to be significant (p < 0.01, Supplementary Table S10), while the CoNVaQ tool resulted in ten significant CNVRs (p < 0.01, Supplementary Table S11). However, no CNVR survived permutation analysis or correction for multiple testing.

### Rare and deleterious single nucleotide variants

A total of 22,703 rare and deleterious variants from 8,442 genes (out of 22,480 genic regions scanned) met criteria for gene burden analysis. On average, cases and controls had 739 ± 51, and 735 ± 52 rare and deleterious variants per sample, respectively (p = 0.779). We identified 114 and 44 genes (p < 0.05) using CMC and SKAT-O, respectively and selected 42 common genes for prioritization analysis (Supplementary Figure S4). Out of 42 genes, 15 genes had a rare and deleterious variant in our cohort (Table 2). All genes survive permutation analysis with *MLLT6* as the top candidate (p = 4.2 × 10^−3^) but no gene survived multiple testing correction. However, compared to the frequencies of rare and deleterious variants in the gnomAD cohort, cases had significantly greater frequencies for seven (*MLLT6, CADPS, TACC2, L3MBTL4, NOL8, FAM172A, AFDN*) of the 15 candidate genes, after Bonferroni correction (Table 2). Frequencies in controls did not differ from those observed in the gnomAD cohort for any of the 15 genes.

**Table 2.** Top hits from gene burden analysis

| Table 2. Top hits from gene burden analysis |  |  |  |  |  |  |  |  |  |
| --- | --- | --- | --- | --- | --- | --- | --- | --- | --- |
| Genes |  |  |  | Case vs Control |  |  | Two Proportion Z-test |  | RVIS<br>(Percentile)<sup>1</sup> |
|  | Frequency<br>Controls<br>(n=24) | Frequency<br>Cases<br>(n=25) | Frequency<br>gnomAD<br>(n=15,708) | CMC<br>P-value | SKAT-O<br>P-value | Empirical<br>P-value | Case vs<br>gnomAD<br>P-value | Control vs<br>gnomAD<br>P-value |  |
| MLLT6 | 0.000 | 0.140 | 0.007 | 0.0051 | 0.0324 | 0.0042 | 3.9x10<sup>-15</sup>* | 0.6818 | -1.51 (6.0%) |
| CADPS | 0.000 | 0.120 | 0.025 | 0.0104 | 0.0207 | 0.0101 | 0.0024* | 0.4354 | -2.30 (2.2%) |
| TACC2 | 0.021 | 0.160 | 0.024 | 0.0119 | 0.0500 | 0.0103 | 9.9x10<sup>-6</sup>* | 0.9203 | 1.92 (96.5%) |
| L3MBTL4 | 0.000 | 0.100 | 0.007 | 0.0208 | 0.0354 | 0.0198 | 3.5x10<sup>-8</sup>* | 0.6818 | 0.38 (66.9%) |
| NPY | 0.000 | 0.100 | 0.032 | 0.0208 | 0.0208 | 0.0208 | 0.0539 | 0.3735 | -0.08 (45.5%) |
| SLC25A21 | 0.021 | 0.140 | 0.037 | 0.0240 | 0.0344 | 0.0209 | 0.0065 | 0.6745 | 0.51 (72.2%) |
| PARVB | 0.000 | 0.100 | 0.025 | 0.0208 | 0.0445 | 0.0211 | 0.0166 | 0.4354 | 0.29 (62.8%) |
| GPR179 | 0.000 | 0.100 | 0.039 | 0.0208 | 0.0399 | 0.0218 | 0.1158 | 0.3222 | 2.80 (98.9%) |
| ACAD9 | 0.000 | 0.100 | 0.028 | 0.0208 | 0.0491 | 0.0218 | 0.0295 | 0.4056 | -0.69 (21.3%) |
| NOL8 | 0.000 | 0.100 | 0.049 | 0.0208 | 0.0491 | 0.0221 | 0.2382 | 0.2670 | 1.92 (96.5%) |
| NADK2 | 0.000 | 0.100 | 0.015 | 0.0208 | 0.0491 | 0.0222 | 0.0005* | 0.5485 | 0.30 (63.4%) |
| FAM172A | 0.021 | 0.140 | 0.016 | 0.0240 | 0.0457 | 0.0225 | 9.2x10<sup>-7</sup>* | 0.8145 | -0.38 (32.2%) |
| AFDN | 0.000 | 0.100 | 0.017 | 0.0208 | 0.0208 | 0.0227 | 0.0013* | 0.5222 | -1.65 (5.0%) |
| SLC6A11 | 0.000 | 0.120 | 0.029 | 0.0208 | 0.0292 | 0.0239 | 0.0068 | 0.3953 | -0.71 (20.7%) |
| PXDN | 0.000 | 0.100 | 0.009 | 0.0208 | 0.0399 | 0.0253 | 0.0348 | 0.6384 | -1.08 (11.4%) |

Comparing frequency of qualifying rare and deleterious variants per gene between cases and controls in the PROCLAIM cohort. Frequencies of the qualifying rare variants in cases and controls were also compared with public controls (gnomAD v2 public database). \*significant after Bonferroni correction (0.05/15 tests = 0.0033). 1 = RVIS Score, a gene-based gene intolerance score based on ExAC v2 release. Genes with negative scores are more “intolerant” than those with positive scores. *MLLT6*: PHD Finger Containing, *CADPS*: Calcium Dependent Secretion Activator, *TACC2*: Transforming Acidic Coiled-Coil Containing Protein 2, *L3MBTL4*: L3MBTL Histone Methyl-Lysine Binding Protein 4, *NPY*: Neuropeptide Y, *SLC25A21*: Solute Carrier Family 25 Member 21, *PARVB*: Parvin Beta, *GPR179*: G Protein-Coupled Receptor 179, *ACAD9*: Acyl-CoA Dehydrogenase Family Member 9, *NOL8*: Nucleolar Protein 8, *NADK2*: NAD Kinase 2, Mitochondrial, *FAM172A*: Family With Sequence Similarity 172 Member A, *AFDN*: Afadin, Adherens Junction Formation Factor, *SLC6A11*: Solute Carrier Family 6 Member 11, *PXDN*: Peroxidas

Gene intolerance analysis showed *CADPS, AFDN*, and *MLLT6* were the most intolerant to rare and deleterious variation, respectively ranking amongst the 2^nd^, 5^th^, and 6^th^ percentile of the most intolerant genes. Gene enrichment analysis of the 15 nominally significant genes revealed no significant functional category or pathway. Likewise, gene enrichment analysis including of the 15 nominally significant genes and the four candidate genes from our previous GWAS (i.e, *SIX3, EFHC1, ADAM7, GNA15*) revealed no significant functional category or pathway. However, all 15 genes, with the exception of *GPR179* and *SLC6A11*, were expressed in left ventricular septal tissue from healthy human adult donors with *TACC2* and *MLLT6* having the highest expression (Figure 3). Among the GWAS candidates, only *SIX3, EFHC1*, and *GNA15* were expressed in left ventricular septal tissue (Supplementary Figure S5).

**Figure 3.**
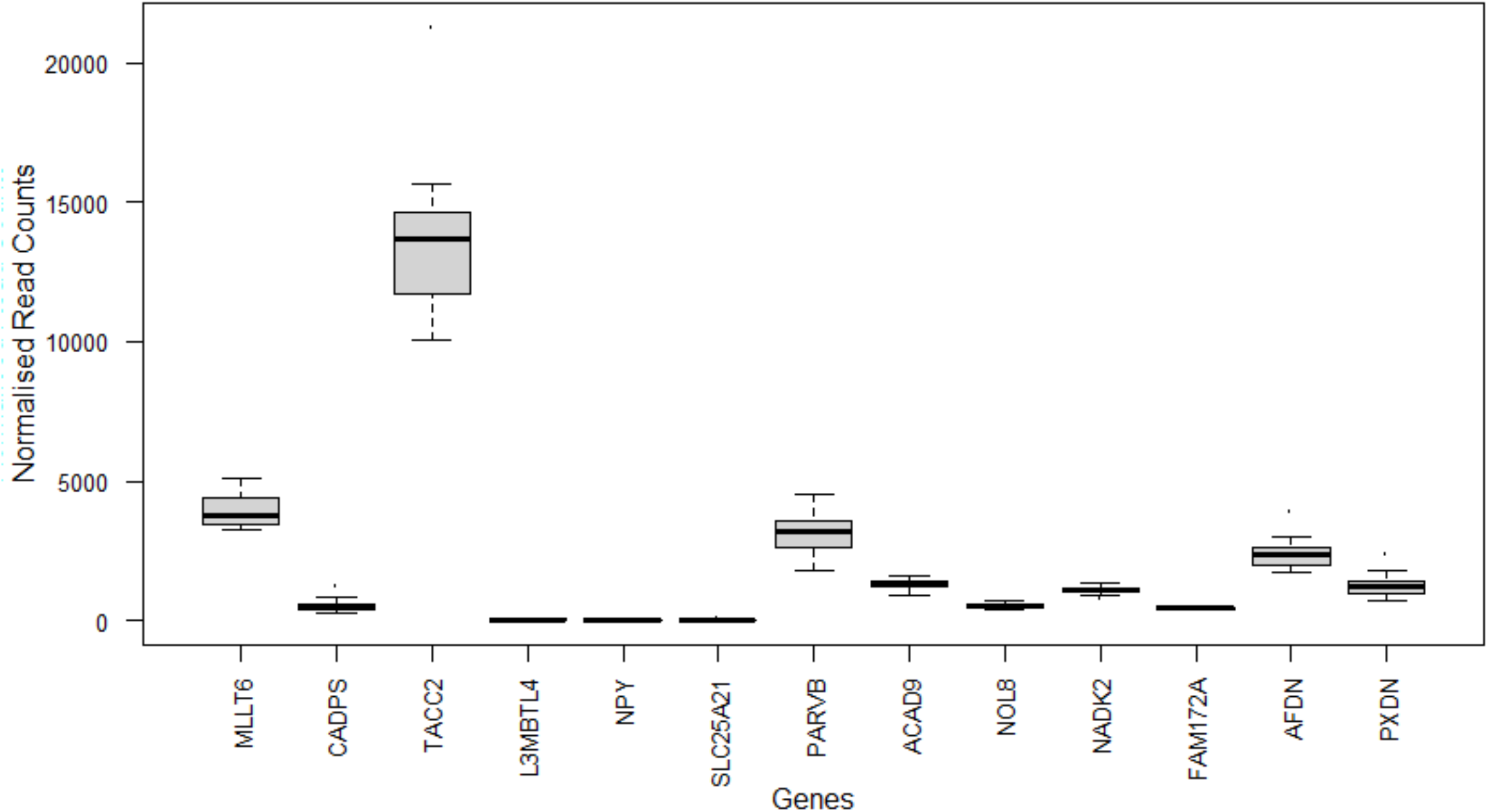
Normalised RNASeq read counts of the 15 nominally associated loci identified in gene burden analysis in left ventricular septal tissue of healthy adult donors (n=21). *MLLT6*: PHD Finger Containing, *CADPS*: Calcium Dependent Secretion Activator, *TACC2*: Transforming Acidic Coiled-Coil Containing Protein 2, *L3MBTL4*: L3MBTL Histone Methyl-Lysine Binding Protein 4, *NPY*: Neuropeptide Y, *SLC25A21*: Solute Carrier Family 25 Member 21, *PARVB*: Parvin Beta, *GPR179*: G Protein-Coupled Receptor 179, *ACAD9*: Acyl-CoA Dehydrogenase Family Member 9, *NOL8*: Nucleolar Protein 8, *NADK2*: NAD Kinase 2, Mitochondrial, *FAM172A*: Family With Sequence Similarity 172 Member A, *AFDN*: Afadin, Adherens Junction Formation Factor, *SLC6A11*: Solute Carrier Family 6 Member 11, *PXDN*: Peroxidasin

## DISCUSSION

Using a whole-genome sequencing approach, our study sought to identify associations between three different and unexplored classes of DNA variation (pharmacogenetic, CNVs, rare variants) and clozapine-induced myocarditis^8^. Although we did not identify a single robust genetic marker for differentiating cases from controls, we identified 15 novel genes nominally associated with this important and sometimes fatal ADR - which continues to limit the use of clozapine, a highly effective medication for schizophrenia. Among the novel candidate genes identified, 13 were found to be expressed in human myocardium, specifically the ventricular septum, providing biological plausibility of a link between these genes and clozapine-induced myocarditis. Although the mechanisms by which these genes may increase risk for clozapine-induced myocarditis remain unclear and require further study, including replication in a separate patient cohort, the genes identified now represent novel targets to inform future functional and clinical studies. Such studies may subsequently improve our understanding of the role played by genetic risk of this serious ADR

Our findings add to the list of candidate genes identified in our previous GWAS (*GNA15, ADAM7, SIX3, EFHC1*) and HLA (*HLA-C*07:01*) studies of clozapine-induced myocarditis^8^ and suggests that the genetic architecture of this serious ADR is complex and likely polygenic. In fact, each of the candidate genes identified to date have low sensitivity and positive predictive value, limiting their clinical utility. This notion aligns with genomic studies of clozapine-induced agranulocytosis, another rare clozapine ADR, that has been associated with variants from several genes with modest sensitivity (range: 11% - 36%)^36^. This current state of the evidence coupled with low incidence of clozapine-induced myocarditis highlights the need for international collaboration to facilitate the rapid ascertainment of cases that will ultimately provide the statistical power required to unravel its complex genetic etiology. Recognizing this need, we formed the PROCLAIM Consortium in 2017 that currently comprises 15 recruitment sites across four countries (Canada, USA, Australia, Turkey)^9^ and are actively looking for new sites to join the consortium.

Our gene enrichment analysis was inconclusive and *post hoc* exploration of the 15 genes using the published literature showed limited to no associations with phenotypes related to myocarditis, inflammation, or adverse drug reactions with few exceptions (see Supplementary Discussion). However, this is not unsurprising given the lack of previously published genetic studies examining this ADR. One notable exception was neuropeptide Y (NPY), which has a well-established role in modulating cardiac contractibility, protein degradation, and proliferation of cardiomyocytes and has been implicated in the pathogenesis of various cardiovascular morbidities (e.g., cardiac hypertrophy, arrhythmias, heart failure)^37^. Furthermore, clinical associations between *NPY* genetic variation and antipsychotic-induced weight gain have been reported^38^. Weight gain and related increases in body mass index (BMI) related to antipsychotic treatment could be clinically relevant given the number of antipsychotic trials patients undergo prior to commencing clozapine and previous work showing high BMI increases clozapine-induced myocarditis fatal outcomes^6, 39^. Notably, BMI did not statistically differ between our cases and controls, although there was a trend (p = 0.107, Cohen’s d = 0.42) toward higher BMIs in cases. Given the paucity of genetic markers for clozapine-induced myocarditis, future investigation of rare genetic variation in *NPY* and the other 14 putative candidate genes should be considered.

Our findings should be interpreted in the context of notable caveats. Since clozapine-induced myocarditis is a relatively rare event - even despite our international consortium - our patient sample size is small and had suboptimal statistical power to detect small-moderate effect sizes. This was partially mitigated by our demographically-matched case-control design and the deployment of an analysis plan appropriate for small sample sizes. Nevertheless, our results are preliminary and will require independent replication before firm conclusions can be drawn. In addition, functional characterization of the identified genes was limited to existing gene expression data derived from healthy adult human left ventricular septal tissue due to the limited biological material and clinical data available from our study participants. Notably, the PROCLAIM Consortium is currently developing a novel *in vitro* model using patient-derived induced pluripotent stem cells differentiated into beating cardiomyocytes to understand how clozapine induces myocardial inflammation and to investigate the impact common and rare genetic variants and/or epigenetic modifications have on cardiomyocyte structure and function when exposed to clozapine.

In summary, our whole-genome sequencing analysis identified 15 genes nominally associated with clozapine-induced myocarditis that provide novel candidates for future studies. With the possible exception of *NPY*, the mechanism by which these genes increase risk for myocarditis is unclear. Although these candidate genes will require independent validation and their roles in the development of clozapine-induced myocarditis remain to be tested, they provide potential insights into the genomic underpinnings and mechanisms by which clozapine induces myocardial inflammation and damage, that may help improve clinical management in the future.

## Data Availability

The data that support the findings of this study are available from the corresponding author, upon reasonable request.

## Acknowledgements

The work was supported in part by the University of Calgary Cumming School of Medicine, Alberta Children’s Hospital Research Institute, and University of Melbourne Establishment Grant. P.L is supported by a National Heart Foundation Future Leader Fellowship (102604).

## Author Contributions

CB and CP conceptualized the study. AN and CB conducted the analyses and wrote the first draft of the manuscript. PL, KR, JM, MJ, NT, RS, DC, RS, SG, and CP contributed to subsequent drafts of the manuscript. All authors approved the final manuscript.

