## Supplementary Material for "Whole-genome sequencing analysis of clozapine-induced myocarditis"

**Supplementary Table S1.** Concordance between Stargazer and

PharmacoScan diplotypes and phenotypes (N=22) …………………………………………………………. 2

**Supplementary Table S2**. List of inhibitors and inducers by

enzyme based on the Flockhart Drug Interactions Table ………………………………………………… 2

**Supplementary Table S3.** Ambiguous CNV regions

excluded from constructing CNVRs ………………………………………………………………………………… 3

**Supplementary Table S4**. WGS statistics for PROCLAIM cohort ……………………………………… 3

### Supplementary Table S5. Observed diplotype frequencies, uncorrected

### activity scores and predicted phenotypes……………………………………………………………………….. 4

### Supplementary Table S6. Uncorrected and corrected activity scores by group………………. 6

### Supplementary Table S7. Uncorrected and corrected metabolizer status by group……..… 8

**Supplementary** **Table S8.** Correlations between activity score

and titration slope…………………………………………………………………………………………………………… 9

**Supplementary Table S9.** Comparison of Number of CNVs

and CNV size between cases and controls ……………………………………………………………………… 10

**Supplementary Table S10.** Significantly CNVRS identified

by CNVRuler in the PROCLAIM Cohort …………………………………………………………………………… 10

**Supplementary Table S11.** Significantly CNVRS identified

by CoNVaQ in the PROCLAIM Cohort …………………………………………………………………………….. 10

**Supplementary Figure S1**. Population structure identified

by principal component analysis ……………………………………………………………………………………. 11

**Supplementary Figure S2**. Distribution of Titration slopes by ancestry………………………….. 12

**Supplementary Figure S3.** Distributions of depth of coverage (top)

and percentage of mapped reads for all 49 samples………………………………………………………. 13

**Supplementary Figure S4.** Overlap in gene burden analysis

conducted using CMC and SKAT-O…………………………………………………………………………………. 14

**Supplementary Figure S5.** Normalised RNASeq read counts in left

ventricular septal tissue of healthy adult donors (n=21) for the four

GWAS loci associated with clozapine-induced myocarditis…………………………………………….. 15

**Supplementary Methods** ……………………………………………………………………………………………… 16

**Supplementary Discussion** …………………………………………………………………………………………… 16

| **Supplementary Table S1.** Concordance between Stargazer and PharmacoScan diplotypes and phenotypes (N=22) | | |
| --- | --- | --- |
| **Genes** | **Diplotype Concordance** | **Phenotype Concordance** |
| CYP2D6 | 18/22 (81.8%) | 22/22 (100%) |
| CYP2C9 | 20/22 (90.9%) | 21/22 (95.5%) |
| CYP2C19 | 21/22 (95.5%) | 21/22 (95.5%) |
| CYP1A2 | 22/22 (100%) | 22/22 (100%) |
| CYP3A4 | 21/22 (95.5%) | 22/22 (100%) |
| CYP3A5 | 22/22 (100%) | 22/22 (100%) |
| CYP2E1 | 22/22 (100%) | 21/22 (95.5%)) |
| **Total** | **146/154 (94.8%)** | **151/154 (98.1%)** |

**Supplementary Table S2**. List of inhibitors and inducers by enzyme based on the Flockhart Drug Interactions Table (https://drug-interactions.medicine.iu.edu/Main-Table.aspx). m=moderate, s=strong.

|  | CYP1A2 | N  Cases/controls | CYP2C19 | N  Cases/controls | CYP2D6 | N  Cases/controls |
| --- | --- | --- | --- | --- | --- | --- |
| Inhibitors | fluvoxamine (s)  ciprofloxacin (s) | 0/0  0/0 | fluoxetine (m)  esomeprazole (m)  ethinylestradiol (m)  voriconazole (m) | 1/0  1/2  0/0  0/0 | fluoxetine (s)  bupropion (s)  paroxetine (s)  es/citalopram (m)  levomepromazine (m)  sertraline (m)  duloxetine (m) | 1/0  0/0  1/0  1/5  0/0  2/1  0/0 |
| Inducers | smoking  carbamazepine  rifampcin  phenytoin  phenobarbital | 6/10  0/0  0/0  0/0  0/0 | carbamazepine  St john’s wort | 0/1  0/0 |  |  |

| **Supplementary Table S3.** Ambiguous CNV regions excluded from constructing CNVRs | | | | |
| --- | --- | --- | --- | --- |
| Sample | Group | Chr | Start | End |
| CIM040 | control | chr1 | 144987033 | 144999817 |
| CIM029 | case | chr10 | 42357882 | 42370804 |
| CIM052 | case | chr12 | 103336058 | 103375908 |
| CIM052 | case | chr15 | 37354672 | 37422581 |
| CIM052 | case | chr16 | 79597944 | 79649705 |
| CIM052 | case | chr17 | 5609392 | 5638927 |
| CIM048 | case | chr21 | 15244028 | 15309588 |
| CIM024 | case | chr7 | 100553479 | 100615535 |
| CIM026 | control | chr9 | 68421016 | 68997842 |
| CIM060 | case | chr9 | 68421016 | 68997842 |

| **Supplementary Table S4**. WGS statistics for PROCLAIM cohort | | |
| --- | --- | --- |
|  | **Mean/Average Statistics** | **Range** |
| SNPs | 3833700.204 | 3789597-3907812 (80.08%) |
| Indels | 953459.5 | 942592-971301 (19.914%) |
| CNVs | 619.18 | 558-670 |
| Number of Duplicate Reads | 11.43% | 6.23-18.72 |
| Alignment | 92.99% | 83.2-95.09 |
| Depth of Coverage | 33.65x | 27.3-36.43 |

### Supplementary Table S5. Observed diplotype frequencies, uncorrected activity scores

### and predicted phenotypes

| Diplotype | Case  Frequency, n (%)^a^ | Control Frequency, n (%)^a^ | Activity Score | Predicted Phenotype^b^ |
| --- | --- | --- | --- | --- |
| *CYP1A2* |  |  |  |  |
| *1F/*1F | 11 (44) | 8 (33) | 3 | UM |
| *1A/*1F | 11 (44) | 10 (42) | 2.5 | UM |
| *1A/*1A | 1 (4) | 1 (4) | 2 | NM |
| *1A/*1L | 0 (0) | 3 (13) | IND | IND |
| *1F/*1L | 1 (4) | 2 (8) | IND | IND |
| *1L/*1L | 1 (4) | 0 (0) | IND | IND |
| *CYP2D6* |  |  |  |  |
| **1/*1x2* | 0 (0) | 1 (4) | 3 | UM |
| **2/*2x2* | 0 (0) | 1 (4) | 3 | UM |
| **2X2/*35* | 1 (4) | 0 (0) | 3 | UM |
| *1/*1 | 4 (16) | 3 (13) | 2 | NM |
| *1/*2 | 4 (16) | 2 (8) | 2 | NM |
| *1/*33 | 2 (8) | 1 (4) | 2 | NM |
| *1/*35 | 1 (4) | 1 (4) | 2 | NM |
| *1/*41 | 1 (4) | 1 (4) | 1.5 | NM |
| *2/*41 | 1 (4) | 1 (4) | 1.5 | NM |
| *1/*36+*10 | 1 (4) | 1 (4) | 1.25 | NM |
| *2/*10 | 1 (4) | 0 (0) | 1.25 | NM |
| *2/*36+*10 | 0 (0) | 1 (4) | 1.25 | NM |
| *1/*4 | 3 (12) | 1 (4) | 1 | IM |
| *2/*4 | 1 (4) | 1 (4) | 1 | IM |
| *2/*4N+*4 | 1 (4) | 1 (4) | 1 | IM |
| *2/*5 | 0 (0) | 1 (4) | 1 | IM |
| *2/*31 | 1 (4) | 0 (0) | 1 | IM |
| *2/*68+*4 | 0 (0) | 1 (4) | 1 | IM |
| *3/*35 | 0 (0) | 1 (4) | 1 | IM |
| *10/*36+*10 | 0 (0) | 1 (4) | 0.5 | IM |
| *4/*41 | 1 (4) | 0 (0) | 0.5 | IM |
| *41/*68+*4 | 0 (0) | 1 (4) | 0.5 | IM |
| *3/*3 | 0 (0) | 1 (4) | 0 | PM |
| *68+*4/68+*4 | 0 (0) | 1 (4) | 0 | PM |
| *41/*122 | 0 (0) | 1 (4) | IND | IND |
| *41/*71 | 2 (8) | 0 (0) | IND | IND |
| *CYP2C19* |  |  |  |  |
| *17/*17 | 1 (4) | 0 (0) | 3 | UM |
| *1/*17 | 5 (20) | 7 (29) | 2.5 | RM |
| *1/*1 | 11 (44) | 8 (33) | 2 | NM |
| *2/*17 | 3 (12) | 1 (4) | 1.5 | IM |
| *1/*2 | 5 (20) | 3 (13) | 1 | IM |
| *2/*2 | 0 (0) | 3 (13) | 0 | PM |
| *2/*3 | 0 (0) | 1 (4) | 0 | PM |
| *2/*8 | 0 (0) | 1 (4) | 0 | PM |
| CYP2C9 |  |  |  |  |
| *1/*1 | 18 (72) | 18 (67) | 2 | NM |
| *1/*2 | 5 (20) | 3 (13) | 1.5 | IM |
| *1/*3 | 2 (8) | 3 (13) | 1 | IM |
| *2/*11 | 0 (0) | 1 (4) | 1 | IM |
| *2/*3 | 0 (0) | 1 (4) | 0.5 | PM |
| CYP2E1 |  |  |  |  |
| *1x2/*7 | 3 (12) | 3 (13) | 2.5 | RM |
| *1/*1 | 16 (64) | 12 (50) | 2 | NM |
| *1/*7x2 | 1 (4) | 0 (0) | 2 | NM |
| *1/*7 | 5 (20) | 9 (38) | 1.5 | IM |
| CYP3A4 |  |  |  |  |
| *1/*1 | 22 (88) | 23 (96) | 2 | NM |
| *1/*22 | 1 (4) | 0 (0) | 1.5 | IM |
| *1/*1B | 1 (4) | 1 (4) | IND | IND |
| *1/*18 | 1 (4) | 0 (0) | IND | IND |
| CYP3A5 |  |  |  |  |
| *1/*1 | 0 (0) | 1 (4) | 2 | NM |
| *1/*3 | 3 (12) | 5 (21) | 1 | IM |
| *3/*3 | 22 (88) | 18 (75) | 0 | PM |

**^a^** Percentages may not equate to 100 due to rounding. ^b^ Predicted phenotypes for *CYP1A2* and *CYP2C19* are based on definitions provided by the Clinical Pharmacogenetics Implementation Consortium (CPIC). CYP2D6 phenotypes were based on the standardized activity score definitions developed jointly by CPIC and the Dutch Pharmacogenetics Working Group. Phenotypes for all other genes were taken directly from Stargazer. IM=intermediate metabolizer, NM=normal (extensive) metabolizer, PM=poor metabolizer, RM=rapid metabolizer, UM=ultrarapid metabolizer, IND=indeterminant.

| **Supplementary Table S6.** Uncorrected and corrected activity scores by group | | | | | | | | |
| --- | --- | --- | --- | --- | --- | --- | --- | --- |
| **Activity Score** | **Group** | **N** | **Mean** | **Median** | **SD** | **SE** | **p** | **Cohen's d** |
| CYP1A2 AS | Case | 23 | 2.72 | 2.50 | 0.30 | 0.06 | 0.721 | 0.11 |
|  | Control | 19 | 2.68 | 2.50 | 0.30 | 0.07 |  |  |
| CYP1A2 PCS | Case | 23 | 3.74 | 3.75 | 0.71 | 0.15 | 1.000 | 0.05 |
|  | Control | 19 | 3.70 | 3.75 | 0.82 | 0.19 |  |  |
| CYP2C19 AS | Case | 25 | 1.88 | 2.00 | 0.56 | 0.11 | 0.574 | 0.38 |
|  | Control | 24 | 1.58 | 2.00 | 0.95 | 0.19 |  |  |
| CYP2C19 PCS | Case | 25 | 1.81 | 2.00 | 0.60 | 0.12 | 0.548 | 0.29 |
|  | Control | 24 | 1.56 | 2.00 | 1.04 | 0.21 |  |  |
| CYP2C9 AS | Case | 25 | 1.86 | 2.00 | 0.23 | 0.05 | 0.576 | 0.24 |
|  | Control | 24 | 1.79 | 2.00 | 0.33 | 0.07 |  |  |
| CYP2C9 PCS | Case | 25 | 1.86 | 2.00 | 0.23 | 0.05 | 0.743 | 0.08 |
|  | Control | 24 | 1.83 | 2.00 | 0.41 | 0.08 |  |  |
| CYP2D6 AS | Case | 23 | 1.61 | 2.00 | 0.58 | 0.12 | 0.331 | 0.27 |
|  | Control | 23 | 1.41 | 1.25 | 0.80 | 0.17 |  |  |
| CYP2D6 PCS | Case | 23 | 1.35 | 1.25 | 0.71 | 0.15 | 0.285 | 0.21 |
|  | Control | 23 | 1.19 | 1.00 | 0.82 | 0.17 |  |  |
| CYP3A4 AS | Case | 23 | 1.98 | 2.00 | 0.10 | 0.02 | 0.339 | -0.29 |
|  | Control | 23 | 2.00 | 2.00 | 0.00 | 0.00 |  |  |
| CYP3A4 PCS | Case | 23 | 1.98 | 2.00 | 0.10 | 0.02 | 0.171 | -0.40 |
|  | Control | 23 | 2.04 | 2.00 | 0.21 | 0.04 |  |  |
| CYP3A5 AS ^a^ | Case | 25 | 0.12 | 0.00 | 0.33 | 0.07 | 0.234 | -0.38 |
|  | Control | 24 | 0.29 | 0.00 | 0.55 | 0.11 |  |  |
| CYP2E1 AS ^a^ | Case | 25 | 1.96 | 2.00 | 0.29 | 0.06 | 0.310 | 0.27 |
|  | Control | 24 | 1.88 | 2.00 | 0.34 | 0.07 |  |  |

AS = Activity score (uncorrected), PCS = phenoconversion-corrected activity score

^a^ Uncorrected and corrected activity scores were identical so only the uncorrected scores are shown.

| **Supplementary Table S7.** Uncorrected and corrected metabolizer status by group | | | | | | |
| --- | --- | --- | --- | --- | --- | --- |
| **Gene** | **Uncorrected, n (%)** | |  | **Corrected, n (%)** | |  |
| **Metabolizer phenotype** | **Case** | **Control** | **p** | **Case** | **Control** | **p** |
| CYP1A2 |  |  | 0.435 |  |  | 0.237 |
| PM | 0 (0) | 0 (0) |  | 0 (0) | 0 (0) |  |
| NM | 1 (4) | 1 (4) |  | 0 (0) | 1 (4) |  |
| UM | 22 (88) | 18 (75) |  | 23 (92) | 18 (75) |  |
| Indeterminant | 2 (8) | 5 (21) |  | 2 (8) | 5 (21) |  |
| CYP2C19 |  |  | 0.087 |  |  | 0.152 |
| PM | 0 (0) | 5 (21) |  | 0 (0) | 5 (21) |  |
| IM | 8 (32) | 4 (17) |  | 9 (36) | 5 (21) |  |
| NM | 11 (44) | 8 (33) |  | 11 (44) | 8 (33) |  |
| RM | 5 (20) | 7 (29) |  | 4 (16) | 5 (21( |  |
| UM | 1 (4) | 0 (0) |  | 1 (4) | 1 (4) |  |
| CYP2C9 |  |  | 0.686 |  |  | 0.517 |
| PM | 0 (0) | 0 (0) |  | 0 (0) | 0 (0) |  |
| IM | 7 (28) | 8 (33) |  | 7 (28) | 8 (33) |  |
| NM | 18 (72) | 16 (67) |  | 18 (72) | 15 (63) |  |
| UM | 0 (0) | 0 (0) |  | 0 (0) | 1 (4) |  |
| CYP2D6 |  |  | 0.504 |  |  | 0.742 |
| PM | 0 (0) | 2 (8) |  | 2 (8) | 2 (8) |  |
| IM | 7 (28) | 8 (33) |  | 9 (36) | 12 (50) |  |
| NM | 15 (60) | 11 (46) |  | 11 (44) | 7 (29) |  |
| UM | 1 (4) | 2 (8) |  | 1 (4) | 2 (8) |  |
| Indeterminant | 2 (8) | 1 (4) |  | 2 (8) | 1 (4) |  |
| CYP3A4 |  |  | 0.513 |  |  | 0.510 |
| PM | 0 (0) | 0 (0) |  | 0 (0) | 0 (0) |  |
| IM | 1 (4) | 0 (0) |  | 1 (4) | 0 (0) |  |
| NM | 22 (88) | 23 (96) |  | 22 (88) | 22 (92) |  |
| UM | 0 (0) | 0 (0) |  | 0 (0) | 1 (4) |  |
| Indeterminant | 2 (8) | 1 (4) |  | 2 (8) | 1 (4) |  |
| CYP3A5 ^a^ |  |  | 0.391 |  |  |  |
| PM | 22 (88) | 18 (75) |  |  |  |  |
| IM | 3 (12) | 5 (21) |  |  |  |  |
| NM | 0 (0) | 1 (4) |  |  |  |  |
| UM | 0 (0) | 0 (0) |  |  |  |  |
| CYP2E1 ^a^ |  |  | 0.371 |  |  |  |
| PM | 0 (0) | 0 (0) |  |  |  |  |
| IM | 5 (20) | 9 (38) |  |  |  |  |
| NM | 17 (68) | 12 (50) |  |  |  |  |
| RM | 3 (12) | 3 (13) |  |  |  |  |

PM = poor metabolizer, IM = intermediate metabolizer, NM = normal metabolizer, RM = rapid metabolizer, UM = ultrarapid metabolizer.

^a^ Uncorrected and corrected metabolizer status frequencies were identical so only the uncorrected metabolizer status frequencies are shown.

**Table S8.** Correlations between activity score and titration slope

| **Activity Score** | **Titration slope** | |
| --- | --- | --- |
|  | **Spearman’s rho** | **p** |
| CYP1A2 AS | 0.14 | 0.389 |
| CYP1A2 PCS | 0.08 | 0.617 |
| CYP2C19 AS | 0.09 | 0.600 |
| CYP2C19 PCS | 0.06 | 0.735 |
| CYP2C9 AS | 0.02 | 0.916 |
| CYP2C9 PCS | 0.02 | 0.916 |
| CYP2D6 AS | -0.11 | 0.525 |
| CYP2D6 PCS | 0.06 | 0.716 |
| CYP3A4 AS | 0.09 | 0.603 |
| CYP3A4 PCS | 0.09 | 0.603 |
| CYP3A5 AS^a^ | -0.16 | 0.339 |
| CYP2E1 AS^a^ | 0.13 | 0.445 |

AS = Activity score (uncorrected), PCS = phenoconversion-corrected activity score

^a^ Uncorrected and corrected activity scores were identical so only the uncorrected scores are shown.

| **Supplementary Table S9.** Comparison of Number of CNVs and CNV size between cases and controls | | | | | | |
| --- | --- | --- | --- | --- | --- | --- |
|  | Student's t Statistic | | df | | | p |
| Number of CNVs | -0.03 |  | 96.0 |  | 0.97 | |
| Gains | 1.13 |  | 47.0 |  | 0.26 | |
| Losses | -0.81 |  | 47.0 |  | 0.42 | |
| CNV Size | -0.63 |  | 30338 |  | 0.523 | |
| Gains | -0.44 |  | 5818 |  | 0.66 | |
| Losses | 0.04 |  | 24518 |  | 0.97 | |

**Supplementary Table S10.** Significantly CNVRS identified by CNVRuler in the PROCLAIM Cohort

| Chr | Start | End | Size | Genes | N, % (Control) | N, % (Case) | Description | p value | Odds Ratio | Bonferroni | FDR |
| --- | --- | --- | --- | --- | --- | --- | --- | --- | --- | --- | --- |
| 1 | 2.49E+08 | 248636065 | 41581 | OR2T7, OR2T2 | 3, 12.5% | 14, 56% | GAIN | 0.00 | 14.98 | 1 | 1.00 |
| 1 | 2.49E+08 | 248797516 | 57698 | OR2T10, OR2T11 | 3, 12.5% | 12, 48% | LOSS | 0.01 | 8.32 | 1 | 1.00 |
| 4 | 6897138 | 6900199 | 3062 | NA | 21, 87.5% | 11, 44% | LOSS | 0.01 | 0.03 | 1 | 1.00 |
| 10 | 48557611 | 48560934 | 3324 | NA | 10, 41.6% | 2, 8% | LOSS | 0.01 | 0.10 | 1 | 1.00 |
| 1 | 7568838 | 7571840 | 3003 | CAMTA1 | 10, 41.6% | 2, 8% | LOSS | 0.01 | 0.10 | 1 | 1.00 |

**Supplementary Table S11.** Significantly CNVRS identified by CoNVaQ in the PROCLAIM Cohort

| Chr. | Start | End | Size | Gene | Status | Control % | Case % | Raw p value | Empirical p value |
| --- | --- | --- | --- | --- | --- | --- | --- | --- | --- |
| 1 | 2.49E+08 | 2.49E+08 | 8531 | OR2T7 | Gain | 8.33-12.5 | 52 | 0.01 | 0.19 |
| 2 | 91701487 | 91703464 | 1979 | Non-genic | Gain | 25 | 64 | 0.01 | 0.34 |
| 15 | 90049915 | 90053105 | 3192 | LINC00928 | Loss | 25 | 0 | 0.01 | 0.69 |
| 4 | 6897138 | 6900199 | 3063 | Non-genic | Loss | 87.5 | 44 | 0.00 | 0.71 |
| 3 | 1.26E+08 | 1.26E+08 | 2333 | UROC1 | Loss | 4.17 | 40-44 | 0.00 | 0.79 |
| 10 | 48557611 | 48559934 | 2325 | Non-genic | Loss | 41.7 | 8 | 0.01 | 0.79 |
| 9 | 1.32E+08 | 1.32E+08 | 2280 | Non-genic | Loss | 0 | 28 | 0.01 | 0.80 |
| 1 | 7569838 | 7571840 | 2004 | CAMTA1 | Loss | 41.7 | 8 | 0.01 | 0.84 |
| 17 | 209982 | 211470 | 1490 | Non-genic | Loss | 29.2 | 0 | 0.00 | 0.89 |
| 13 | 54237812 | 54238913 | 1103 | Non-genic | Loss | 0 | 28 | 0.01 | 0.94 |

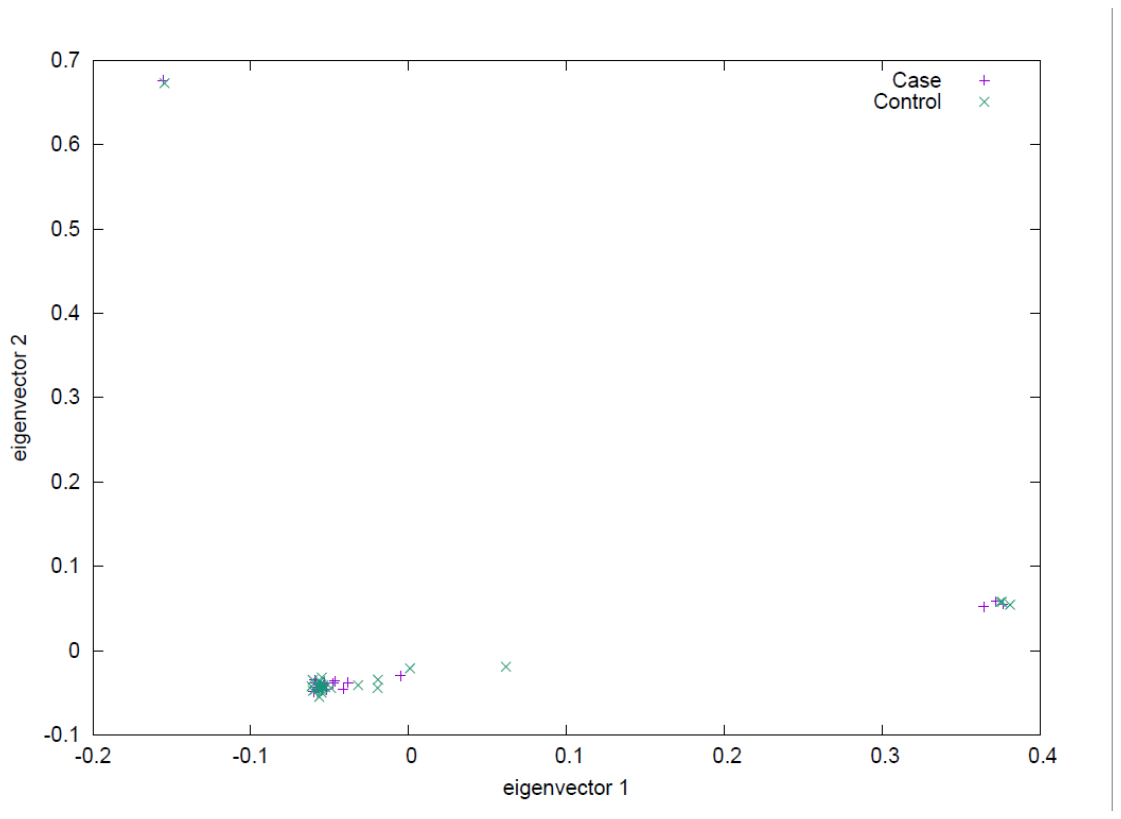

**Supplementary Figure S1**. Population structure identified by principal component analysis. The first two components are plotted against each other. Cases (n=25) are depicted in purple (+) and controls (n=24) are depicted in green (x).

| Pairwise comparisons - Titration Slope | | | | | | |
| --- | --- | --- | --- | --- | --- | --- |
|  | |  | | **W** | | **p** |
| EAS |  | EUR |  | 1.201 |  | 0.396 |
| EAS |  | NEA |  | 0.548 |  | 0.698 |
| EAS |  | OCE |  | 0.414 |  | 0.770 |
| EAS |  | SAS |  | -2.070 |  | 0.143 |
| EUR |  | NEA |  | 0.000 |  | 1.000 |
| EUR |  | OCE |  | -0.316 |  | 0.823 |
| EUR |  | SAS |  | -2.373 |  | 0.093 |
| NEA |  | OCE |  | 0.000 |  | 1.000 |
| NEA |  | SAS |  | -1.732 |  | 0.221 |
| OCE |  | SAS |  | -1.414 |  | 0.317 |

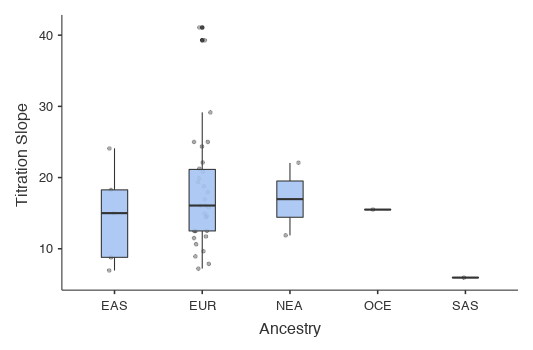

**Supplementary Figure S2**. Distribution of Titration slopes by ancestry. EAS, East Asian; EUR, European; NEA, Near Eastern, OCE, Oceanian; SAS, South Asian

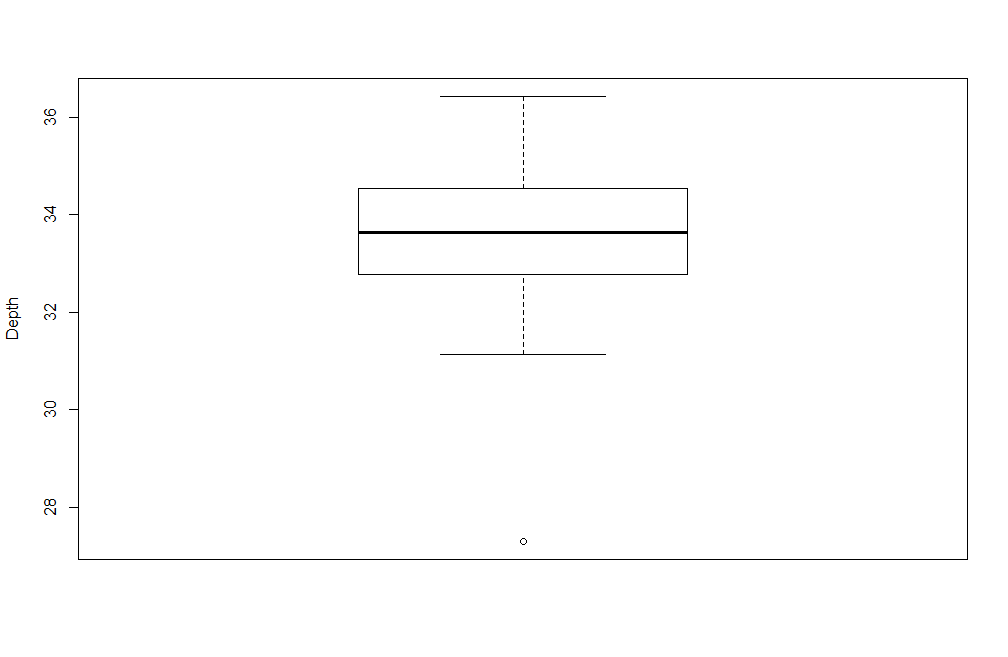

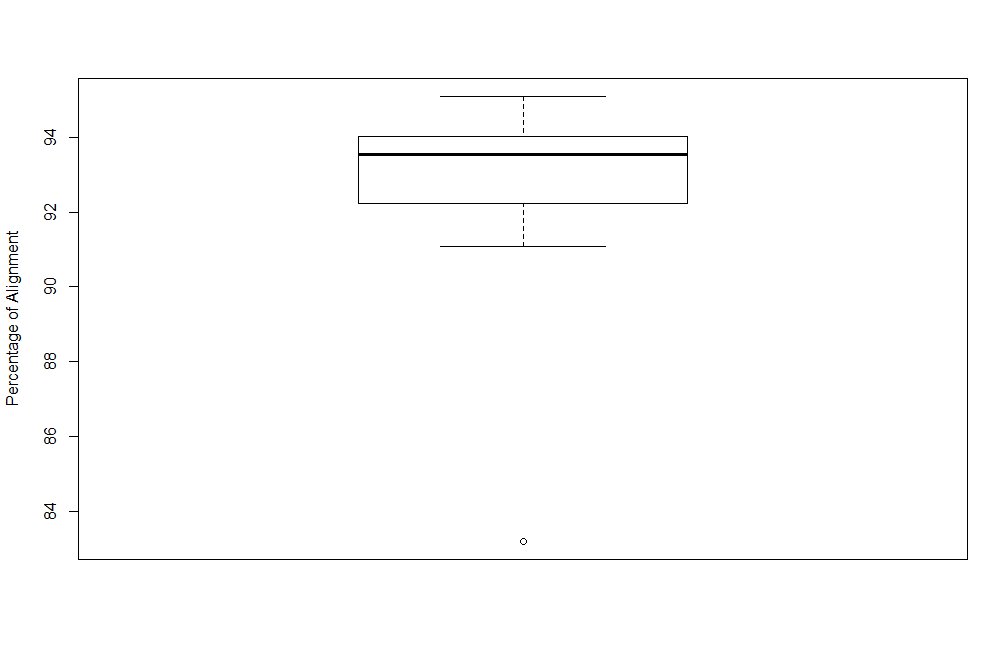

**Supplementary Figure S3.** Distributions of depth of coverage (top) and percentage of mapped reads for all 49 samples.

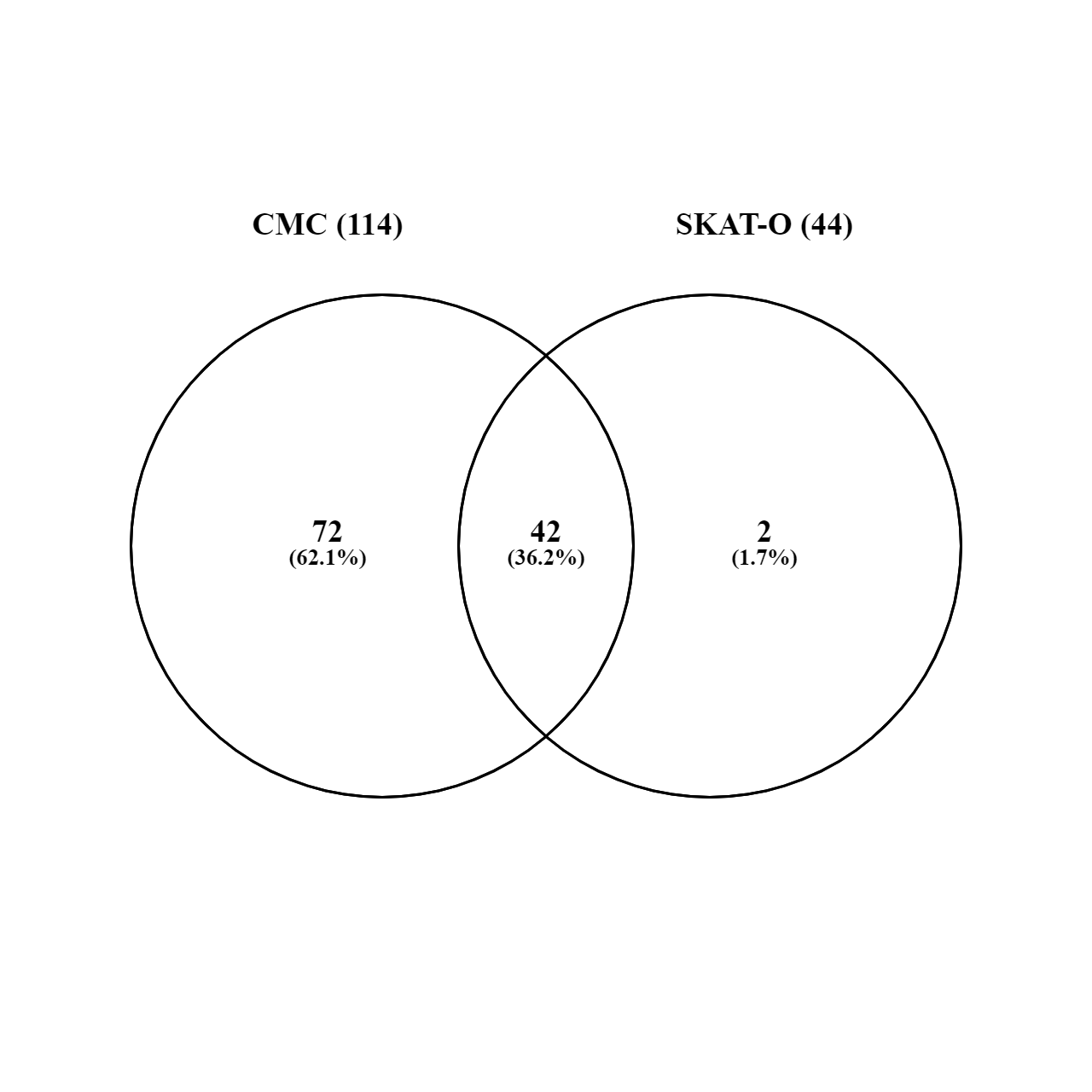

**Supplementary Figure S4.** Overlap in gene burden analysis, conducted using CMC and SKAT-O.

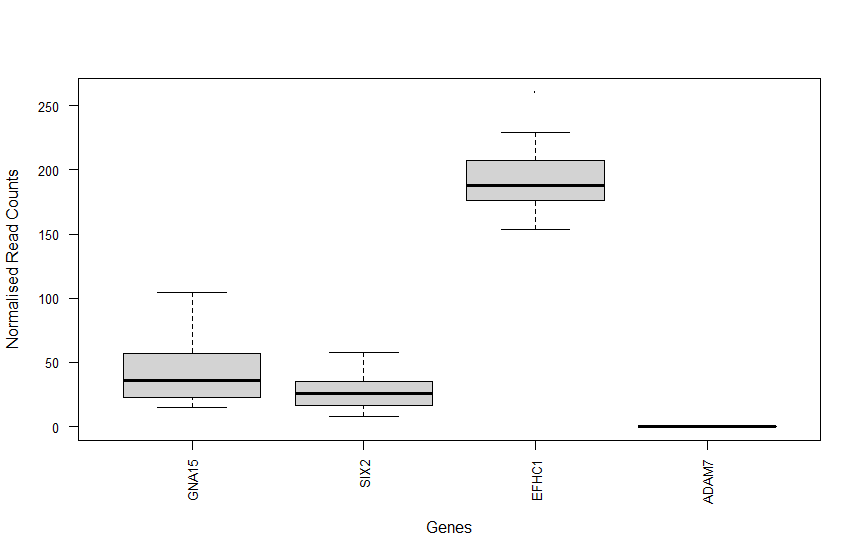

**Supplementary Figure S5.** Normalised RNASeq read counts in left ventricular septal tissue of healthy adult donors (n=21) for the four GWAS loci associated with clozapine-induced myocarditis.

**Supplementary Methods**

Training sites/resources for VQSR algorithm for SNPs and Indels:

**SNPs:**

(i) hapmap_3.3.hg19.sites.vcf
(ii) illumina_platinum.snps.hg19.sites.vcf
(iii) 1000G_omni2.5.hg19.sites.vcf
(iv) 1000G_phase1.snps.high_confidence.hg19.sites.vcf, dbSNP150.vcf.gz

**Indels:**

(i) Mills_and_1000G_gold_standard.indels.hg19.sites.vcf
(ii) illumina_platinum.indels.hg19.sites.vcf

**Supplementary Discussion**

Our three aforementioned mutationally intolerant genes (*CADPS*, *AFDN*, and *MLLT6)* may warrant further scrutiny. *CADPS* encodes CAPS1 (Ca2+-activated protein for secretion-1, also known as CAPS and UNC-31), a cardiac, neural- and endocrine-specific peptide involved in calcium-regulated exocytosis of secretory vesicles, including those containing catecholamines and NPY, specifically in the biogenesis and storage of large dense-core vesicles (LDCVs). Homozygous knockout in mice was reportedly lethal within 30 minutes of birth (possibly due to apnea), while deletion of one allele distorted the intracellular distribution of LDCVs, and decreased their secretion and docking ^1^. *CADPS* is expressed in minor cell types of the murine spleen, where it has been hypothesized that it might therefore be involved in release of neurotrophin-3, brain-derived neurotrophic factor, vasoactive intestinal peptide, and somatostatin ^2^; however, expression in lymphoid tissues is very low, and no clear links to drug-induced or viral myocarditis more generally have been established. Nevertheless, the following (albeit more indirect) lines of evidence do raise the possibility of a pathophysiological association.

In a GWAS study of educational attainment – blood pressure interaction, amongst 18 new genome-wide significant loci the largest effect on systolic BP (12.85 mm Hg) was for rs141962517 a SNP intronic to *CADPS*, of note given a significant positive dose-response relationship between plasma catecholamine levels and severity of myocardial inflammation in a murine model of clozapine-induced myocarditis, attenuated by propranolol ^3^. In colorectal cancer cells, where CAPS1 expression is frequently upregulated, CAPS1 induced epithelial-mesenchymal transition (EMT) ^4^, a process involved in injury recovery and repair, while strain-specific myocarditis-like changes and EMT (the latter interpreted as part of a healing process) have been reported in C57BL/6j mice stressed by aggressive intruder attacks. ^5^

CAPS1 is only secreted in the cardiac atria under healthy conditions, but appears in the ventricles of mice with calcineurin-induced and afterload-induced cardiac hypertrophy. It interacts with RRP17 (Ras-related protein 17), which is encoded by *RASL10B*, and specifically expressed in cardiomyocytes, neurons, and pancreas. RRP17 positively regulates both intracellular CAPS1 levels and secretion by cardiomyocytes of atrial natriuretic peptide (ANP), which regulates blood pressure and urinary sodium excretion. In response to acute and chronic stress, synthesis and secretion of ANP and BNP (brain/B-type natriuretic peptide) is upregulated by ventricular myocytes. Notably, both ANP and BNP bind to the same guanylyl cyclase-linked receptor, natriuretic peptide receptor-A, causing vasodilation, natriuresis, and diuresis, as well as inhibition of endothelin-1 release and the renin–angiotensin system. BNP and the N-terminal fragment of its propeptide (NT-pro-BNP) are commonly used to diagnose and monitor cardiac failure, and NT-pro-BNP was 91% sensitive and 73% specific in identifying patients with left ventricular failure in one study ^6^, although data is too limited to recommend its use for routine monitoring of clozapine cardiotoxicity ^7^. Knockout of *RASL10B*/RRP17 in mice impaired ANP release, and induced hypertension and tachycardia in anaesthetized mice^8^, although it is unclear whether this effect was mediated through abrogation of RRP17-CAPS1 signaling.

Taken together, the aforementioned studies raise the possibility that, given the right genetic background, defective CAPS1 signaling might impair recovery and compensation from myocardial inflammation induced by a stress and/or clozapine-induced hypercatecholaminergic state.

Curiously, the remaining two of our three most variation-intolerant genes: *MLLT6, and AFDN*, also known as *MLLT4*, are implicated in the formation of leukemogenic MLLT (Myeloid/Lymphoid Or Mixed-Lineage Leukemia; Translocated To) fusion proteins formed through chromosomal translocations with truncated forms of *MLL* (*KMT2A*). *MLL/KMT2A* encodes a histone lysine methyltransferase functioning as a transcriptional regulator. The resulting in-frame fusions promote dimerization of the N-terminal region of this gene, result in a gain-of-function of this gene, which is involved in regulating hematopoiesis. As of 2017, at least 94 fusion partners of *KMT2A*/*MLL* had been identified ^9^; to our knowledge, amongst our 15 significant genes, only *AFDN/MLLT4* and *MLLT6* share this association with *MLL*. These two genes are otherwise very dissimilar; they have no domains in common, and subserve very different functions.

*MLLT6*, which encodes the AF17 protein, may be of particular interest, as it has been identified as a transcriptional regulator that positively modulates the expression of the PD-L1 (programmed cell death 1 ligand 1), encoded by *CD274*. Binding of the PD-L1 cell surface protein to the PD1 receptor on T cells inhibits T-cell activation and cytokine productions and suppresses CD8^+^ cytotoxic T cell-mediated cytolysis. High expression levels of *MLLT6* are associated with poor survival in gastric cancer, while knockout resulted in significant reduction of PD-L1, increased CD8^+^ T cell-mediated cytolysis, and reduced expression of interferon-χ-associated immune resistance ^10^. Intriguingly, PD-L1 expression on cardiac myocytes suppressed T cell activation, while inhibition of PD1-PDL1 interaction on T-lymphocytes by the monoclonal antibody nivolumab led to T-lymphocyte activation, and an attendant autoimmune cardiomyopathy associated with increased myocardial TNF-α levels, T-lymphocyte infiltration, and cardiomyocyte apoptosis ^11^. However, while *MLLT6* is fairly ubiquitously expressed at the mRNA level, myocardial expression is low ^12^, and while sHet^13^ predicts heterozygous loss-of-function variants to be deleterious, the gene is not haploinsufficient by another metric ^14^.

*AFDN/MLTT4* encodes afadin (AF6), an actin filament (F-actin) and Rap1 small G protein-binding protein, which has been implicated primarily in cell-cell adhesion at adherens junctions, but it may also play a role in cell polarization, migration, survival, and proliferation, cooperatively regulating formation of cell adhesion together with its binding partner nectin ^15^. Of note, afadin forms adhesion complexes with nectin and c-Cbl-associated protein (CAP) also known as ponsin, which is encoded by the *SORBS1* (Sorbin and SH3 domain-containing protein 1) gene, a member of the SoHo vinexin family of adapter proteins. CAP/ponsin has been shown in a murine model of viral (Coxsackie virus B3) myocarditis to suppress CD8^+^ T lymphocyte and natural killer cell-mediated cytotoxicity, while CAP knockout markedly increased myocardial toxicity and mortality. Conceivably, disruption of afadin-CAP interactions by *AFDN* mutations could have a similar effect. However, it should be noted that (although neutrophilic and lymphocytic cardiac infiltration are also prominent in descriptions of autopsied cases), the mechanism underlying clozapine-induced myocarditis may involve an eosinophilic hypersensitivity reaction^4,5,39,16^, and eosinophilia is not described in association with either *AFDN/MLLT4* or *MLLT6* knockdown.
